# When might Host Heterogeneity Drive the Evolution of Asymptomatic, Pandemic Coronaviruses?

**DOI:** 10.1101/2020.12.19.20248566

**Authors:** Kenichi W. Okamoto, Virakbott Ong, Robert Wallace, Rodrick Wallace, Luis Fernando Chaves

**Affiliations:** Department of Biology, University of St. Thomas, St. Paul MN 55105 USA; Agroecology and Rural Economics Research Corps, St. Paul MN USA; New York State Psychiatric Institute, NYC, NY USA; Institute of Tropical Medicine, Nagasaki University, Nagasaki, 852-8521 Japan and Instituto Costarricense de Investigación y Ensenanza en Nutrición y Salud, San José, Costa Rica

## Abstract

For most emerging infectious diseases, including SARS-Coronavirus-2 (SARS-CoV-2), pharmaceutical intervensions such as drugs and vaccines are not widely available, and disease surveillance followed by isolating, contact-tracing and quarantining infectious individuals is critical for controlling outbreaks. These interventions often begin by identifying symptomatic individuals. However, by actively removing pathogen strains likely to be symptomatic, such interventions may inadvertently select for strains less likely to result in symptomatic infections. Additionally, the pathogen’s fitness landscape is structured around a heterogeneous host pool. In particular, uneven surveillance efforts and distinct transmission risks across host classes can drastically alter selection pressures. Here we explore this interplay between evolution caused by disease control efforts, on the one hand, and host heterogeneity in the efficacy of public health interventions on the other, on the potential for a less symptomatic, but widespread, pathogen to evolve. We use an evolutionary epidemiology model parameterized for SARS-CoV-2, as the widespread potential for silent transmission by asymptomatic hosts has been hypothesized to account, in part, for its rapid global spread. We show that relying on symptoms-driven reporting for disease control ultimately shifts the pathogen’s fitness landscape and can cause pandemics. We find such outcomes result when isolation and quarantine efforts are intense, but insufficient for suppression. We further show that when host removal depends on the prevalence of symptomatic infections, intense isolation efforts can select for the emergence and extensive spread of more asymptomatic strains. The severity of selection pressure on pathogens caused by these interventions likely lies somewhere between the extremes of no intervention and thoroughly successful eradication. Identifying the levels of public health responses that facilitate selection for asymptomatic pathogen strains is therefore critical for calibrating disease suppression and surveillance efforts and for sustainably managing emerging infectious diseases.

## Introduction

Epidemiologically, the 2020 SARS-Coronavirus-2 (SARS-CoV-2) pandemic differs markedly from the 2003 SARS-Coronavirus-1 (SARS-CoV-1) outbreak. In particular, the global scale and incidence of the 2020 pandemic stands in sharp contrast with the largely regional scale of the 2003 SARS epidemic (e.g., Chen et al. 2020b; Mahase 2020; Petersen et al. 2020; Prem et al. 2020; Xu et al. 2020). While the underlying pathogens and human host populations in which the outbreak occurred differ along several biological and social axes, one of the major differences has been the degree to which individuals infected with SARS-CoV-2 are unlikely to present symptoms (Nishiura et al. 2020; Mizumoto et al. 2020; Oran and Topol 2020; but see Lee et al. 2003). This results in fewer cases being detected, and a higher proportion of unwittingly infectious carriers circulating in the population. The large fraction of asymptomatic hosts therefore presents a major epidemiological challenge when facing pathogens such as SARS-CoV-2 (Rahimi and Talebi Bezmin Abadi 2020; Lai et al. 2020; Wilder-Smith et al. 2020).

Simultaneously, the SARS-CoV-2 pandemic of 2020 has reinforced the centrality of non-pharmaceutical interventions to suppress and mitigate outbreaks of emerging zoonotic pathogens (Cheng et al. 2020; Davies et al. 2020). Most emerging zoonotic pathogens such as coronaviruses lack well-tested, widely and immediately available pharmaceutical interventions, especially as many veterinary pharmaceuticals remain untested for human use (Qualls et al. 2017; Bird and Mazet 2018). Thus, the selection pressures these pathogens experience from public health interventions have the potential to be very different from the selection pressures operating on endemic pathogens. For instance, with the important exception of vector-borne diseases, an endemic pathogen’s evolutionary responses to control strategies typically result from interventions on the primary (often human) host. By contrast, emerging zoonotic pathogens must evolve within a fitness landscape characterized mainly by host heterogeneity.

From the perspective of pathogen evolution, a major consequence of host heterogeneity is that the intensity of non-pharmaceutical intervention efforts likely differs across host classes. For instance, disease surveillance efforts among human populations far outpaces efforts in animal, an especially sylvatic, populations (e.g., Keusch et al. 2010; Halliday et al. 2017). As a result, the fitness consequences of public health interventions on pathogens are likely to be experienced differently by pathogens infecting different host types.

Even after zoonotic transmission, host heterogeneity among humans can continue to define the fitness landscape of pathogens (Lo Iacono et al. 2016).For instance, social differences can be a major driver of host heterogeneity in zoonotic disease risk (Dzingirai et al. 2017a,b; Leach et al. 2017). Economic disparities in particular are likely to result not only in distinct transmission and infection risks among hosts, but also in variable public health responses (Levins 1995; Kawachi and Kennedy 1999; Navarro and Shi 2001; Szreter and Woolcock 2004; Wallace et al. 2015). The lack of pharmaceutical interventions, and the heavy reliance on public institutions for disease control may exacerbate existing disparities (Marmot 2005; Waitzkin 2018; Stojkoski et al. 2020). This is because non-pharmaceutical interventions, ranging from highly targeted case-detection and expansive contact-tracing to population-wide guidelines on physical distancing to broad-scale shutdowns, require both a robust public health infrastructure and high social and economic development to implement successfully (Farmer 1996; Jamison DT, Breman JG, Measham AR, et al. 2006; Garrett 2007; Jones et al. 2008; Dye 2014; Whitmee et al. 2015; National Research Council 2016; Birn, Anne-Emanuelle and Holtz 2017; Halliday et al. 2017; Wood et al. 2017; Lim and Sziarto 2020). At its best, disease control relying on isolating and tracking severe cases can ensure that limited public health resources are optimally allocated to reducing transmission (Eames and Keeling 2003; Klinkenberg et al. 2006; Armbruster and Brandeau 2010; Rodrigues et al. 2014; Peak et al. 2017). By July 2020, these policies proved tentatively effective in suppressing SARS-CoV-2 in some cases when carried out with precision, multi-sector societal collaboration and adequate financial and operational support (Lu et al. 2020; Cheng et al. 2020; Watson et al. 2020; Chaves et al. 2020; World Health Organization 2020). By contrast, several jurisdictions have also struggled to contain the outbreak (Campbell and Doshi 2020; Solis et al. 2020; World Health Organization 2020).

Yet we argue that even under generally favorable conditions, intervention measures that fall short of disease eradication could allow strains that are able to circulate with minimal host removal to readily outcompete strains that cause their hosts to be detected. When asymptomatic carriers are not detected by the epidemiological monitoring system, the pathogens they harbor are able to spread with less friction through the host population. By contrast, potentially symptomatic lineages are subject to detection and even asymptomatic, exposed contacts are quarantined, thereby pruning that particular viral lineage. We wish to be clear. Our goal here is not to argue against the value of case detection at the point of care and subsequent, vigorous contact tracing. Rather, our point is that symptoms-based surveillance is a necessary, but, from an evolutionary perspective, potentially insufficient component of a robust public health strategy aimed at preventing disease emergence. Furthermore, the adequacy of any broad-scale non-pharmaceutical intervention is likely to be compromised by differential attention and resources aimed at different host groups - be they sylvatic or human, economically prosperous or neglected, or otherwise. If reducing disease burden is a long-term objective, localized containment, particularly if it is haphazardly implemented across host classes, may prove to be evolutionarily unsustainable.

We interrogate this possibility using a mathematical model of a population of hosts in which multiple coronavirus strains, some more symptomatic than others, circulate. Superficially, selection for a largely asymptomatic strain may seem like a tolerable epidemiological outcome. However, we argue that high prevalence of undetected infections is a major reason for why host heterogeneity is important. As the 2020 SARS-CoV-2 pandemic cruelly illustrates, widespread circulation of a generally asymptomatic pathogen raises risks for vulnerable host subpopulations in which the infection is unlikely to be mild (Borjas 2020; Lippi et al. 2020; Chen et al. 2020a; Wu et al. 2020; Li et al. 2020), and there is growing evidence of differences among host populations driving the efficacy of public health interventions and transmission risks (Abedi et al. 2020; Stojkoski et al. 2020; Vahidy et al. 2020). Hence, we model the evolutionary dynamics of multiple strains of a pathogen circulating among heterogeneous hosts consisting of two host types: hosts vulnerable to infections and experiencing limited public health attention, and hosts less likely to be infected and experiencing greater public health attention. Because host heterogeneity plays a critical role in structuring the fitness landscapes of emerging zoonotic pathogens, we seek to elucidate how such heterogeneity interacts with the selective effects of non-pharmaceutical intervention strategies on pathogen evolution, particularly as it concerns selection for disease severity. Below, we present the structure of our model and our analyses.

## Materials and Methods

### Model development

We consider a pathogen circulating within a heterogeneous host population. The host population consists of two main host classes: vulnerable hosts, which experience potentially higher disease transmission but are also potentially less likely to be subject to disease surveillance, and resilient hosts, who have reduced infection risk and are potentially likelier to be the target of disease surveillance. We focus our analyses on the case where the individual transition of hosts between the two host classes over the course of their lifetime is negligible (as may occur, for instance, with limited socioeconomic mobility) or nonexistent (as in the case of sylvatic and human hosts).

## A Baseline Model without Interventions

We assume that transmission is frequency-dependent. Thus, if a susceptible host of type *h* encounters infectious hosts of type *h′*, pathogen transmission occurs at a per-capita rate *βr*_*h,h*′_*/N*, where *N* is the density of all susceptible and infectious hosts, *β* is the baseline per-capita infection rate, and *r*_*h,h*′_ describes the extent of mixing between hosts *h* and *h′*. Following infection, we assume individuals of host type *h* infected with strain *j* become asymptomatic with strain-specific probability *π*_*j,h,A*_. During infection, strain *i* may emerge via mutation, and subsequently out-compete strain *j* within a host at a per-infected individual rate *µ*_*j→i*_ (e.g., Saloniemi 1993; Bohannan and Lenski 2000; Rice 2004). For brevity, we refer to this process by which an infected individual of strain *j* undergoes a within-host replacement by strain *i* as “mutation”. Furthermore, because of the relatively low case fatality rate of SARS-CoV-2 (e.g., Russell et al. 2020; Mizumoto and Chowell 2020), we consider infection-induced mortality to have minimal epidemiological effects over the time scales of interest. Similarly, the incubation period is treated as being sufficiently short to not affect evolutionary trajectories.

Finally, infectious individuals recover from the infection at rate *γ*. In the absence of evidence of antagonistic pleiotropy, we treat this rate as consistent across strains. Furthermore, for SARS-coronoviruses, there is as yet no robust, widely available vaccine and limited evidence of lifelong immunity (Galanti and Shaman 2020; Kellam and Barclay 2020; Edridge et al. 2020). Should these conditions change, we think our model will remain relevant as coronaviruses are subject to repeated and regular zoonotic emergences and re-emergences (e.g., Lee and Hsueh 2020) for which there is unlikely to be pre-existing immunity. Thus, we consider the period of complete immunity over evolutionary-epidemiological timescales to be relatively short, at least as it concerns the evolutionary dynamics of symptomatic and less-symptomatic strains. Taking all of the above into consideration, in the absence of any public-health interventions (pharmaceutical or otherwise), the resulting evolutionary epidemiology can be summarized as:

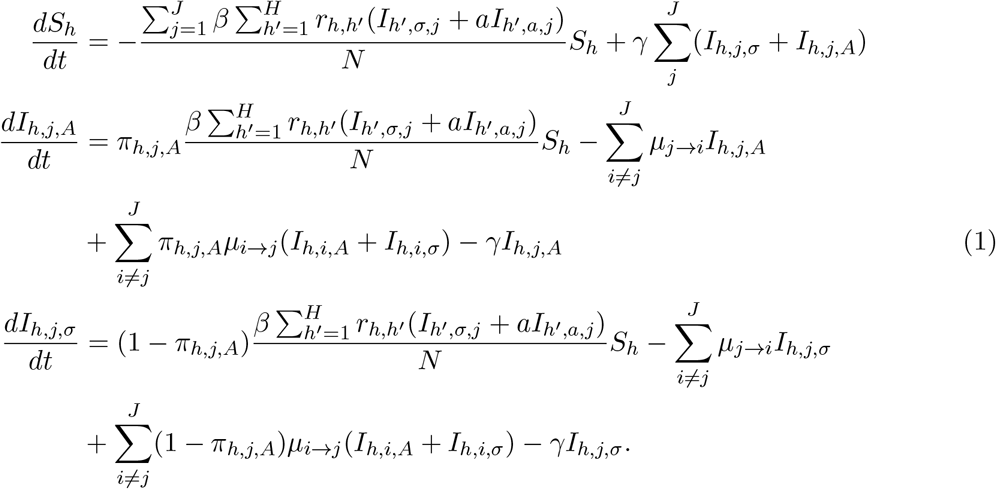

Table 1 summarizes the parameter values of model (1).

**Table 1.**
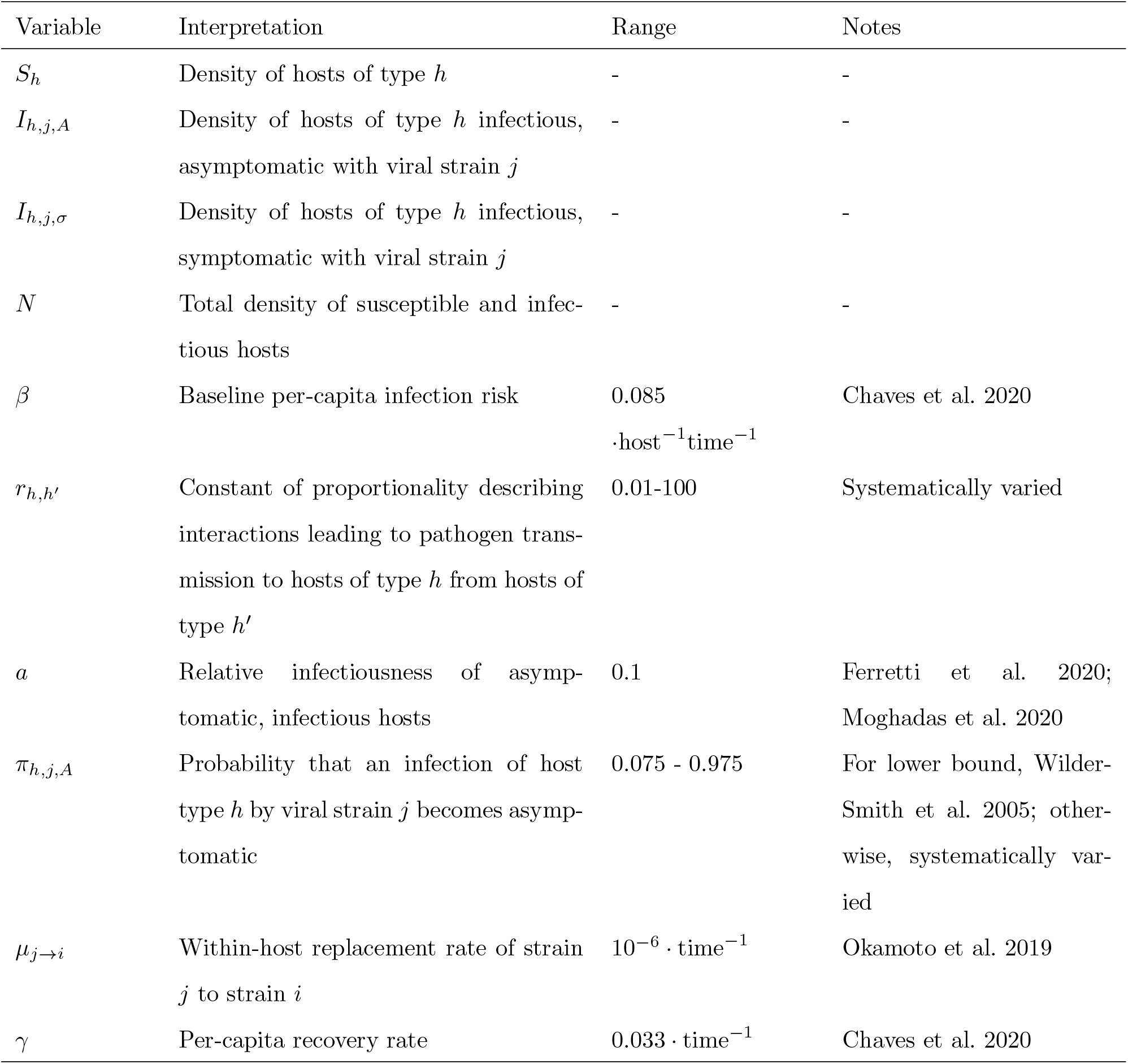
Variables for the baseline model without any interventions.

| Variable | Interpretation | Range | Notes |
| --- | --- | --- | --- |
| $S_h$ | Density of hosts of type $h$ | - | - |
| $I_{h,j,A}$ | Density of hosts of type $h$ infectious, asymptomatic with viral strain $j$ | - | - |
| $I_{h,j,\sigma}$ | Density of hosts of type $h$ infectious, symptomatic with viral strain $j$ | - | - |
| $N$ | Total density of susceptible and infectious hosts | - | - |
| $\beta$ | Baseline per-capita infection risk | 0.085<br>$\cdot \text{host}^{-1} \text{time}^{-1}$ | Chaves et al. 2020 |
| $r_{h,h'}$ | Constant of proportionality describing interactions leading to pathogen transmission to hosts of type $h$ from hosts of type $h'$ | 0.01-100 | Systematically varied |
| $a$ | Relative infectiousness of asymptomatic, infectious hosts | 0.1 | Ferretti et al. 2020; Moghadas et al. 2020 |
| $\pi_{h,j,A}$ | Probability that an infection of host type $h$ by viral strain $j$ becomes asymptomatic | 0.075 - 0.975 | For lower bound, Wilder-Smith et al. 2005; otherwise, systematically varied |
| $\mu_{j \rightarrow i}$ | Within-host replacement rate of strain $j$ to strain $i$ | $10^{-6} \cdot \text{time}^{-1}$ | Okamoto et al. 2019 |
| $\gamma$ | Per-capita recovery rate | $0.033 \cdot \text{time}^{-1}$ | Chaves et al. 2020 |

### The effect of public health interventions

To model the effect of public health interventions as well as their potential unequal application across host classes on the evolution of viral phenotypes, we focus on two types of interventions currently used. First, we assume that symptomatic individuals can be successfully identified and isolated at a potentially time-varying, per-capita rate *θ*_*h,j*_(*t*) that depends on the host type *h* and the strain type *j* with which they are infected. Symptomatic individuals that are isolated are removed from the population of infectious hosts. Because isolated hosts eventually recirculate in the population after recovery, the density of isolated hosts is denoted by *K*_*h,j*_ for isolated hosts of type *h* infectious with strain *j*.

Second, we model the effect of subsequent contact tracing and testing aimed at identifying asymptomatic carriers. We assume that this contact tracing and testing can identify infected, asymptomatic hosts of type *h* infected with strain *j* at a potentially time-varying, per-capita rate *q*_*h,j*_(*t*). Once asymptomatic carriers are identified through contact tracing and testing, they are also removed from the population of infectious hosts. As with isolated, symptomatic hosts, these asymptomatic cases are ultimately returned to the population after the infection clears, so we track the density *Q*_*h,j*_ of successfully removed, asymptomatic hosts of type *h* carrying strain *j*. To distinguish the removal of asymptomatic and symptomatic hosts, we refer to the removal of asymptomatic hosts following contact tracing and testing as “quarantining”, and the removal of symptomatic hosts as “isolation”. We use *v*_*q*_, *v*_*θ*_ to describe the relative difference in quarantining and isolation efficacy between the two strains (*q*_*h,j*_(*t*)*/q*_*h,j′*_ (*t*) and *θ*_*h,j′*_(*t*)*/θ*_*h,j′*_ (*t*), respectively). Successfully quarantined or isolated hosts do not come into contact with susceptible and infectious individuals; thus, their densities do not contribute to *N*, which, as noted above, we define as the sum of infectious and susceptible hosts. We further treat the testing regime during the decision to isolate or quarantine as sufficiently effective (e.g., Yates et al. 2020; Watson et al. 2020) that the accidental removal of uninfected hosts (i.e., false positives) is negligible.

Successfully isolated or quarantined hosts recover at an accelerated rate *δ* that represents, for instance, the effects of administering antiviral drugs (Wang et al. 2020) or the successful management of symptoms (Horby et al. 2020). Because asymptomatic individuals may, potentially, be subject to ongoing monitoring, we treat the enhanced per-capita recovery rate of quarantined individuals as comparable to the enhanced per-capita recovery rate of isolated individuals.

The combined eco-evolutionary dynamics of the interacting hosts and pathogens in the presence of a public health response are given by:

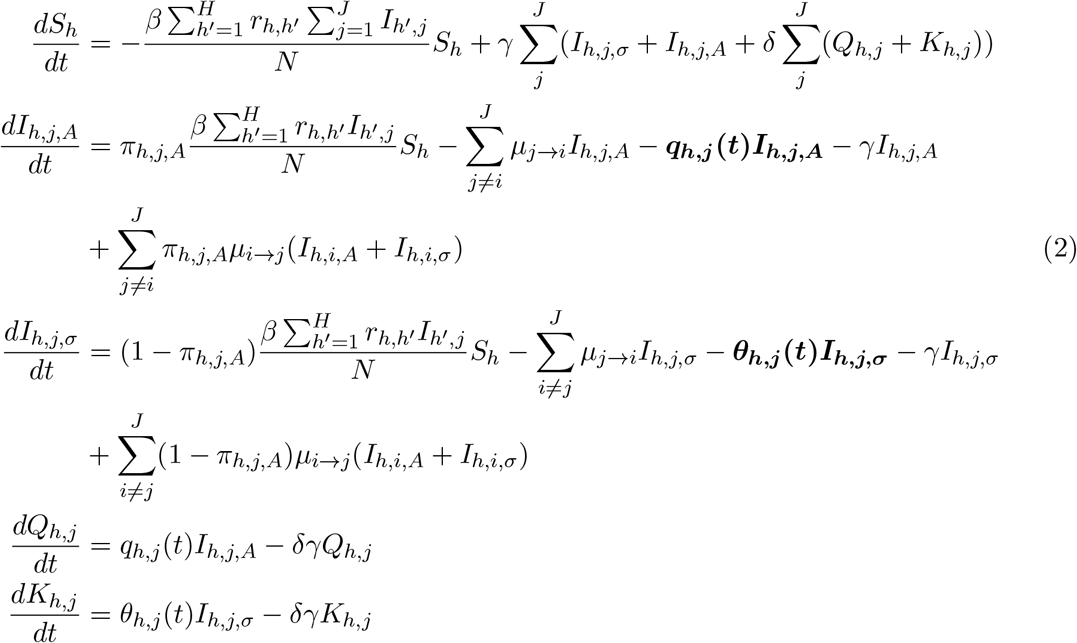

for pathogen strains *j* = 1, …, *J* and host types *h* = 1, …, *H*. We highlight two further points regarding model (2). First, because successfully quarantined and isolated hosts do not contribute to onward pathogen transmission, we ignore mutation dynamics within successfully isolated or quarantined hosts as those viral mutants cannot spread further. Second, we do not model the effects of public health policies that aim to reduce the supply of susceptible hosts (e.g., mass lock-downs, culling, etc…). This is because we seek to model how interventions based on surveillance in the pre-pandemic stage select for different pathogen strains. Thus, here we concern ourselves with evolution that occurs before large-scale public health interventions reducing the density of susceptibles become necessary. Nevertheless, we hasten to add that our model does allow for public health measures aimed at transmission reduction (e.g., by providing access to personal protective equipment): the composite term *β*_*j,h*_*r*_*h,h*′_ governing transmission is able to account for such effects. Table 2 summarizes the key parameters added to model (1) in model (2).

**Table 2.** Additional variables used to model public health interventions.

| Variable | Interpretation | Range |
| --- | --- | --- |
| $Q_{h,j}$ | Density of quarantined hosts of type $h$ infected with viral strain $j$ | - |
| $K_{h,j}$ | Density of isolated hosts of type $h$ infected with viral strain $j$ | - |
| $q_{h,j}(t)$ | The efficacy of quarantining asymptomatic infections of hosts of type $h$ by viruses of type $j$ | 0.0001-1 |
| $\theta_{h,j}(t)$ | The efficacy of isolating asymptomatic infections of hosts of type $h$ by viruses of type $j$ | 0.0001-1 |
| $v.$ | The relative ability to successfully identify and remove symptomatic and asymptomatic infections involving the novel virus | 0.0001-1 |
| $\delta$ | The accelerated recovery of quarantined or isolated hosts | 0.05-0.85 |

### Model Analyses

We explore how public health interventions and differences between host classes drive the evolutionary emergence of asymptomatic strains of SARS-coronaviruses. We focus on two scenarios. We begin by examining the scenario where an ongoing, background surveillance effort leads to generally constant, host-class and virus-strain specific quarantining and isolation efforts over time (i.e., *q*_*h,j*_(*t*) *≈ q*_*h,j*_ and *θ*_*h*_(*t*) *≈ θ*_*h,j*_). We then assess the effects of reactive surveillance efforts, in which the quarantining and isolation efforts depend on the prevalence of symptomatic infections 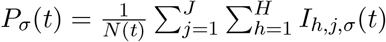. Because infection prevalence is time-varying, under this scenario, *q*_*h,j*_(*t*) = *q*_*h,j*_*P* (*t*) and *θ*_*h,j*_(*t*) = *θ*_*h,j*_*P* (*t*). We assume that, over the evolutionary timescales we consider, the ability of public health responses to respond to prevalence is effectively instantaneous.

Because of the high-dimensionality of model (2), we focus our analyses on numerical explorations; even the relatively simpler baseline model (1) did not lend itself to ready algebraic or analytic characterization. Tables 1-2 summarize the numerical range of parameter values used. Additionally, to facilitate biological interpretation, we present results for two pathogen strains - an ancestral strain and a novel, less-symptomatic strain, systematically varying the extent to which infection by the novel strain is likely to be asymptomatic. All numerical analyses are conducted using the function lsoda from the package deSolve in R (Soetaert et al. 2010), with absolute error tolerance set at 10^*-*50^. We initialized our analyses with a total host population density of 10^6^ individuals per unit area, with a ten-to-one ratio of vulnerable to resilient hosts. Epidemics in all analyses are seeded with a single infectious individual with the ancestral, symptomatic strain. All code used in the analysis is accessible on github (https://github.com/kewok/AsymptomaticEvolution) and is released under the GNU Public License v3 (Stallman 2007).

## Results

We highlight results showing how the efficacy of public health interventions and their heterogeneous application to distinct host classes drives the evolution and spread of asymptomatic coronaviruses. In the context of public health, the prevalence of infection by the novel strain is of greater concern than the frequency of the asymptomatic virus in the viral population *per se*. Thus, we illustrate our results using the long-term prevalence of hosts infected with the evolved, asymptomatic strain to characterize the joint evolutionary and epidemiological predictions. In the comparisons that follow, we distinguish between a baseline isolation efficacy and a baseline efficacy of contact-tracing followed by quarantining for the ancestral strain, and the effectiveness of isolation and contact tracing followed by quarantining for the derived, more asymptomatic strain. We also compare the effects of increased infection risk and reduced efficacy for the vulnerable host class.

### Scenario 1: Constant effort for public health interventions

When isolation and contact-tracing followed by quarantine (hereafter “quarantining”) are carried out at a constant rate (*q*_*h,j*_(*t*) = *q*_*h,j*_ and *θ*_*h,j*_(*t*) = *θ*_*h,j*_), a wide range of qualitatively distinct evolutionary outcomes result (Fig. 1). Generally, increasing isolation effort selects for a novel strain to spread in the host population, until the isolation efficacy is sufficiently high that disease control occurs before an asymptomatic mutant can evolve. Exceptions are Fig. 1J and scenarios resulting in complete suppression or prevalences of 100% across the entire parameter space (not shown in Fig 1).

**Figure 1:**
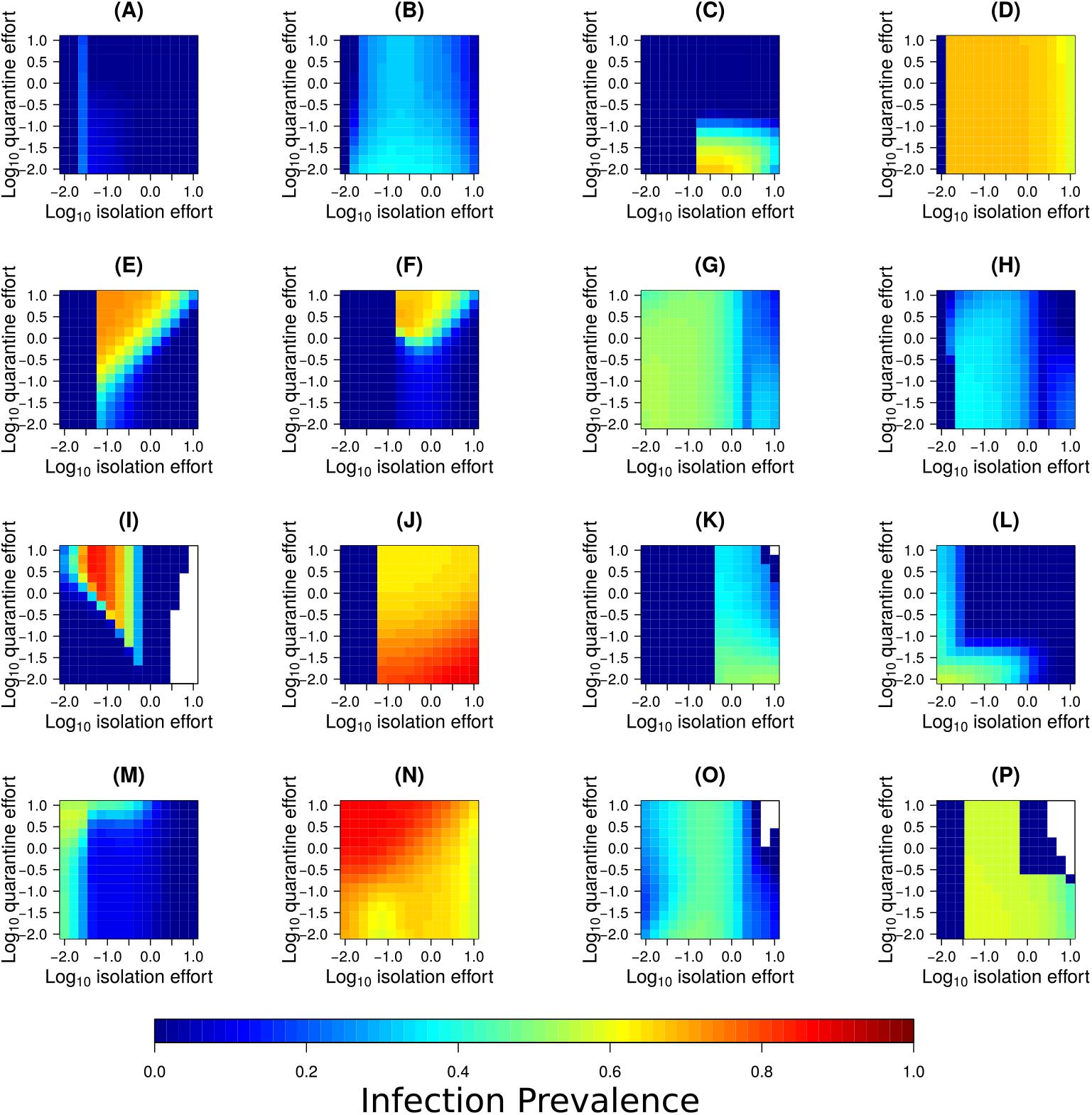
The range of possible qualitative behavior of long-term prevalence of all hosts infected with the novel, more asymptomatic virus in model (2) as a function of quarantine and isolation effort. In addition to the results above, the model also produced outcomes where the novel virus could not successfully spread or would infect all hosts irrespective of the isolation and quarantine efforts (results not shown). (A) *r*_*h,h*′_ *=* 1, *π*_1,2_*/π*_1,1_ = 5, *π*_2,2_*/π*_2,1_ = 13, *v*_*q*_ = *v*_*θ*_ = 0.001 and *q*_*h*,·_*/q*_*h*′,·_ = *θ*_*h*,·_*/θ*_*h*′,*cdot*_ = 1. (B) *r*_*h,h*′_ *=* 1, *π*_1,2_*/π*_1,1_ = 13, *π*_2,2_*/π*_2,1_ = 5, *v*_*q*_ = 1, *v*_*θ*_ = 0.001 and *q*_*h*,·_*/q*_*h*′,·_ = *θ*_*h*,·_*/θ*_*h*′,*cdot*_ = 1. (C) *r*_*h,h*′_ *=* 1, *π*_1,2_*/π*_1,1_ = 5, *π*_2,2_*/π*_2,1_ = 13, *v*_*q*_ = 1, *v*_*θ*_ = 0.001 and *q*_*h*,·_*/q*_*h*′,·_ = *θ*_*h*,·_*/θ*_*h*′,*cdot*_ = 0.1. (D) *r*_*h,h*′_ *=* 10, *π*_1,2_*/π*_1,1_ = 5, *π*_2,2_*/π*_2,1_ = 5, *v*_*q*_ = 0.001, *v*_*θ*_ = 0.001 and *q*_*h*,·_*/q*_*h*′,·_ = *θ*_*h*,·_*/θ*_*h*′,*cdot*_ = 1. (E) *r*_*h,h*′_ *=* 10, *π*_1,2_*/π*_1,1_ = 5, *π*_2,2_*/π*_2,1_ = 13, *v*_*q*_ = 0.001, *v*_*θ*_ = 0.001 and *q*_*h*,·_*/q*_*h*′,·_ = *θ*_*h*,·_*/θ*_*h*′,*cdot*_ = 1. (F) *r*_*h,h*′_ *=* 1, *π*_1,2_*/π*_1,1_ = 5, *π*_2,2_*/π*_2,1_ = 13, *v*_*q*_ = 0.001, *v*_*θ*_ = 0.001 and *q*_*h*,·_*/q*_*h*′,·_ = *θ*_*h*,·_*/θ*_*h*′,*cdot*_ = 0.1. (G) *r*_*h,h*′_ *=* 0.1, *π*_1,2_*/π*_1,1_ = 13, *π*_2,2_*/π*_2,1_ = 1.1, *v*_*q*_ = 0.001, *v*_*θ*_ = 0.001 and *q*_*h*,·_*/q*_*h*′,·_ = *θ*_*h*,·_*/θ*_*h*′,*cdot*_ = 0.1. (H) *r*_*h,h*′_ *=* 0.1, *π*_1,2_*/π*_1,1_ = 13, *π*_2,2_*/π*_2,1_ = 5, *v*_*q*_ = 0.001, *v*_*θ*_ = 0.001 and *q*_*h*,·_*/q*_*h*′,·_ = *θ*_*h*,·_*/θ*_*h*′,*cdot*_ = 0.1. (I) *r*_*h,h*′_ *=* 10, *π*_1,2_*/π*_1,1_ = 1.1, *π*_2,2_*/π*_2,1_ = 1.1, *v*_*q*_ = 0.001, *v*_*θ*_ = 1 and *q*_*h*,·_*/q*_*h*′,·_ = *θ*_*h*,·_*/θ*_*h*′,*cdot*_ = 1. (J) *r*_*h,h*′_ *=* 10, *π*_1,2_*/π*_1,1_ = 5, *π*_2,2_*/π*_2,1_ = 1.1, *v*_*q*_ = 0.001, *v*_*θ*_ = 0.001 and *q*_*h*,·_*/q*_*h*′,·_ = *θ*_*h*,·_*/θ*_*h*′,*cdot*_ = 0.1. (K) *r*_*h,h*′_ *=* 10, *π*_1,2_*/π*_1,1_ = 13, *π*_2,2_*/π*_2,1_ = 5, *v*_*q*_ = 1, *v*_*θ*_ = 0.001 and *q*_*h*,·_*/q*_*h*′,·_ = *θ*_*h*,·_*/θ*_*h*′,*cdot*_ = 0.1. (L) *r*_*h,h*′_ *=* 0.1, *π*_1,2_*/π*_1,1_ = 1.1, *π*_2,2_*/π*_2,1_ = 1.1, *v*_*q*_ = 1, *v*_*θ*_ = 0.001 and *q*_*h*,·_*/q*_*h*′,·_ = *θ*_*h*,·_*/θ*_*h*′,*cdot*_ = 0.1. (M) *r*_*h,h*′_ *=* 0.1, *π*_1,2_*/π*_1,1_ = 1.1, *π*_2,2_*/π*_2,1_ = 13, *v*_*q*_ = 0.001, *v*_*θ*_ = 0.001 and *q*_*h*,·_*/q*_*h*′,·_ = *θ*_*h*,·_*/θ*_*h*′,*cdot*_ = 0.1. (N) *r*_*h,h*′_ *=* 1, *π*_1,2_*/π*_1,1_ = 1.1, *π*_2,2_*/π*_2,1_ = 5, *v*_*q*_ = 0.001, *v*_*θ*_ = 0.001 and *q*_*h*,·_*/q*_*h*′,·_ = *θ*_*h*,·_*/θ*_*h*′,*cdot*_ = 0.1. (O) *r*_*h,h*′_ *=* 0.1, *π*_1,2_*/π*_1,1_ = 13, *π*_2,2_*/π*_2,1_ = 1.1, *v*_*q*_ = 1, *v*_*θ*_ = 0.001 and *q*_*h*,·_*/q*_*h*′,·_ = *θ*_*h*,·_*/θ*_*h*′,*cdot*_ = 0.1. (P) *r*_*h,h*′_ *=* 10, *π*_1,2_*/π*_1,1_ = 13, *π*_2,2_*/π*_2,1_ = 1.1, *v*_*q*_ = 1, *v*_*θ*_ = 0.001 and *q*_*h*,·_*/q*_*h*′,·_ = *θ*_*h*,·_*/θ*_*h*′,*cdot*_ = 1. Here, and for subsequent figures, we note that varying the rate *δ* at which isolated or quarantined hosts recovered had very little effect on long-term prevalence (Supplementary Material S1 and S2).

Across the parameter space we explore, a critical distinguishing feature among the range of dynamical behaviors is whether the interaction between isolation and quarantine effort promotes the evolution and spread of the novel, more asymptomatic virus. The evolutionary consequences of isolation can, in some cases, occur largely irrespective of the quarantine effort (e.g., Figs. 1A, 1D, 1G, 1H, 1O). When quarantining also affects viral evolution and spread, we find increasing quarantine efforts can have divergent effects. At times, increased removal of symptomatic hosts selects for an asymptomatic strain, but this can be mitigated by more effective quarantining. For instance, Figs. 1B-C, 1J-L and 1P show how even at levels of isolation effort selecting for an asymptomatic strain, effective quarantining can mitigate the spread of the evolved virus (cooler colors in upper regions of those panels). By contrast, Figs. 1E-F, 1I and 1M-N illustrate the opposite effect: higher quarantining efforts interact with levels of isolation that select for asymptomatic viruses to drive high prevalence. Even within the latter scenarios, the joint effects of isolation and quarantining are not always consistent. For instance, Figs. 1E-1F and 1N show how the interaction between high quarantine levels and isolation diminishes as isolation efficacy increases.

Supplementary Tables S3 and S4 systematically group all the scenarios we analyzed into each of these qualitative long-term patterns based on direct visual inspection of all parameter combinations. Supplementary Table S3 categorizes the effects for the case where the two host classes receive disparate public health intervention efforts; Supplementary Table S4 evaluates the conditions when the two host classes receive comparable public health intervention efforts.

A comparison of scenarios in Supplementary Table S3 reveals how the ability to detect asymptomatic cases can be a major driver of the distinctions highlighted above. Indeed, the differing effects of increasing quarantine effort arise because, like isolation effort, it is ultimately intermediate levels of quarantining that select for the emergence and spread of an asymptomatic variant. In our model, the net quarantine effort for the novel virus is a composite of the ability to detect asymptomatic hosts infected with either strain, and a baseline quarantine effort. Thus, whether increasing quarantining selects for an asymptomatic virus critically depends upon the ability to detect asymptomatic infections. Comparing Fig. 1C to Fig. 1F illustrates how an ability to detect asymptomatic infections shifts the evolutionary effects of quarantining. When the ability to detect asymptomatic infections is limited (Fig. 1F), higher quarantining efforts promote the evolution and spread of an asymptomatic strain. Yet as the ability to detect asymptomatic infections improves (Fig. 1C), increasing quarantining efforts can successfully suppress the asymptomatic strain. A further driver of the qualitative differences is the transmission risk for the resilient host (Table S3). Because these hosts are likelier to be the target of strong public health interventions, increased transmission in this host class can result in greater selection on the asymptomatic strain, until sufficiently high isolation efforts in this host class facilitate successful disease suppression. Supplementary Table S3 again illustrates how this outcome depends on whether there is a reasonable prospect of identifying asymptomatic mutant infections.

Finally, in addition to public health interventions and host heterogeneities, the probability (*π*_*h,j,A*_) that infections from the evolved strain are asymptomatic also mediates the nature of how quarantine and isolation effort interact to drive the evolution and spread of the asymptomatic virus. For instance, when the ability to detect asymptomatic cases is low, as the evolved virus becomes increasingly asymptomatic (particularly towards the resilient host), the effect of increasing quarantine efforts changes from successful suppression to facilitating the evolution of the asymptomatic strain at intermediate isolation efforts (Supplementary Table S3). This shift occurs because strains that cause more asymptomatic infections are harder to suppress even when quarantine efforts are high, whereas viruses less likely to cause asymptomatic infections are more readily controlled by increasing quarantine efforts. Once more, we see how these effects are magnified when the probability of being asymptomatic is high for resilient hosts, because the resilient host is also likelier to be subject to increased isolation and quarantine efforts.

These results thusfar characterize the evolutionary and epidemiological consequences of host heterogeneities in public health responses. When there is a more even application of isolation and quarantining across host classes (so that *q*_*h,j*_ = *q*_*h′,j*_, *h* ≠ *h′*), a somewhat different pattern emerges. In general, we find that more uniformity in transmission risk and isolation and quarantining efforts between vulnerable and resilient hosts usually reduces the prevalence of the more asymptomatic virus (Fig. 2A-C and 2D-F). An exception is when isolation efforts are very low but disease suppression otherwise results (Fig 2G-I). In this scenario, very modest isolation efforts prevent the evolutionary emergence of the asymptomatic strain when the public health response is unequal (Fig. 2G), but can result in selective pressure for the more asymptomatic strain when public health responses and transmission risk are more even across host classes (Fig. 2I-H).

**Figure 2:**
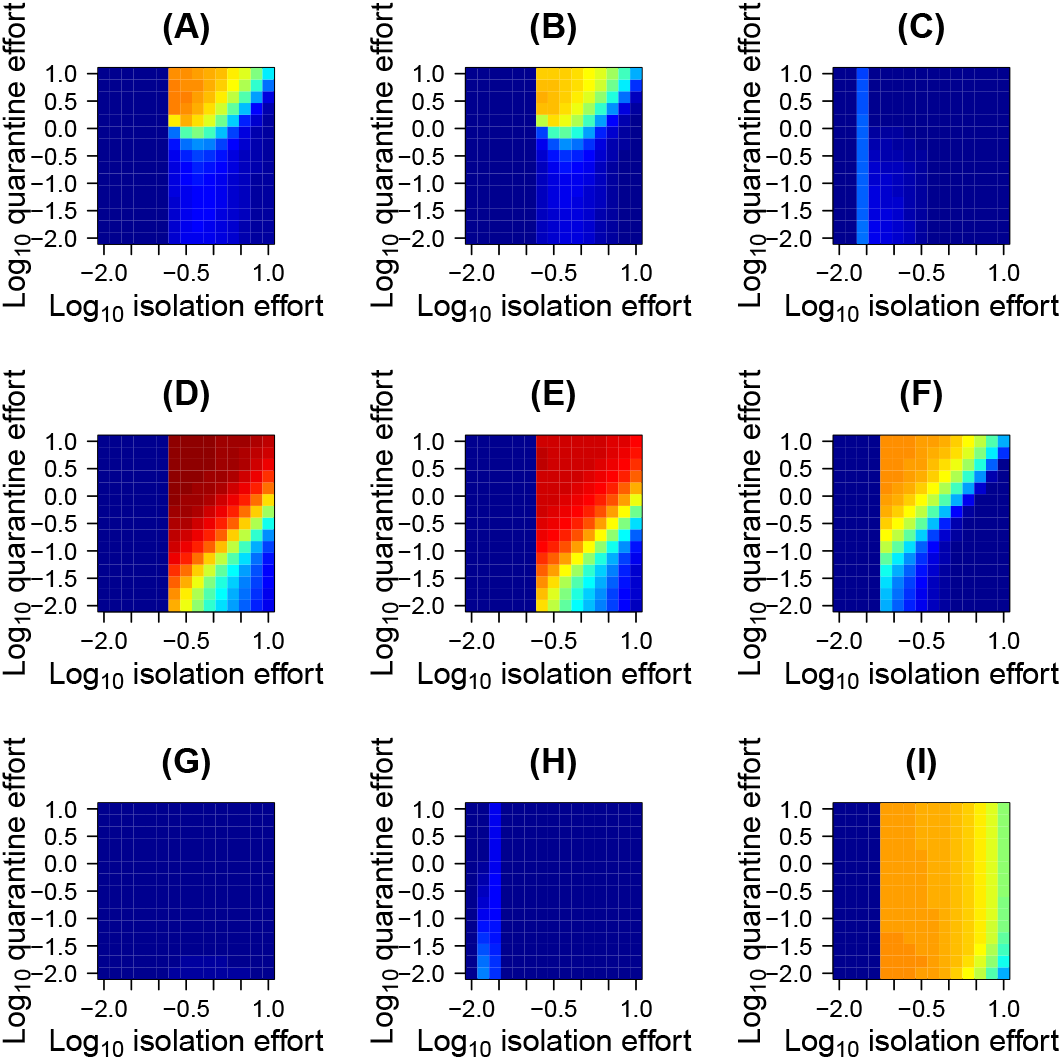
The effect of increasing isolation and quarantine efforts or reducing transmission risk for the neglected host, in terms of long-term prevalence of among hosts infected with the novel, more asymptomatic virus in model (2) as a function of quarantine and isolation effort. In this, and in subsequent figures, the color scheme follows Figure 1. (A) *r*_*h,h*′_ *=* 10, *π*_1,2_*/π*_1,1_ = 13, *π*_2,2_*/π*_2,1_ = 13, *v*_*q*_ = *v*_*θ*_ = 0.001 and *q*_*h*,·_*/q*_*h*′,·_ = *θ*_*h*,·_*/θ*_*h*′,*cdot*_ = 0.1.(B) *r*_*h,h*′_ *=* 1, *π*_1,2_*/π*_1,1_ = 13, *π*_2,2_*/π*_2,1_ = 13, *v*_*q*_ = *v*_*θ*_ = 0.001 and *q*_*h*,·_*/q*_*h*′,·_ = *θ*_*h*,·_*/θ*_*h*′,*cdot*_ = 0.1. (C) *r*_*h,h*′_ *=* 1, *π*_1,2_*/π*_1,1_ = 13, *π*_2,2_*/π*_2,1_ = 13, *v*_*q*_ = *v*_*θ*_ = 0.001 and *q*_*h*,·_*/q*_*h*′,·_ = *θ*_*h*,·_*/θ*_*h*′,*cdot*_ = 1. (D) *r*_*h,h*′_ *=* 100, *π*_1,2_*/π*_1,1_ = 13, *π*_2,2_*/π*_2,1_ = 13, *v*_*q*_ = *v*_*θ*_ = 0.001 and *q*_*h*,·_*/q*_*h*′,·_ = *θ*_*h*,·_*/θ*_*h*′,*cdot*_ = 0.11. (E) *r*_*h,h*′_ *=* 10, *π*_1,2_*/π*_1,1_ = 13, *π*_2,2_*/π*_2,1_ = 13, *v*_*q*_ = *v*_*θ*_ = 0.001 and *q*_*h*,·_*/q*_*h*′,·_ = *θ*_*h*,·_*/θ*_*h*′,*cdot*_ = 1. (F) *r*_*h,h*′_ *=* 10, *π*_1,2_*/π*_1,1_ = 13, *π*_2,2_*/π*_2,1_ = 13, *v*_*q*_ = 0.001, *v*_*θ*_ = 0.001 and *q*_*h*,·_*/q*_*h*′,·_ = *θ*_*h*,·_*/θ*_*h*′,*cdot*_ = 1. (G) *r*_*h,h*′_ *=* 1, *π*_1,2_*/π*_1,1_ = 5, *π*_2,2_*/π*_2,1_ = 13, *v*_*q*_ = 1, *v*_*θ*_ = 0.001 and *q*_*h*,·_*/q*_*h*′,·_ = *θ*_*h*,·_*/θ*_*h*′,*cdot*_ = 0.1. (H) *r*_*h,h*′_ *=* 0.1, *π*_1,2_*/π*_1,1_ = 5, *π*_2,2_*/π*_2,1_ = 13, *v*_*q*_ = 1, *v*_*θ*_ = 0.001 and *q*_*h*,·_*/q*_*h*′,·_ = *θ*_*h*,·_*/θ*_*h*′,*cdot*_ = 1. (I) *r*_*h,h*′_ *=* 10, *π*_1,2_*/π*_1,1_ = 5, *π*_2,2_*/π*_2,1_ = 13, *v*_*q*_ = 1, *v*_*θ*_ = 0.001 and *q*_*h*,·_*/q*_*h*′,·_ = *θ*_*h*,·_*/θ*_*h*′,*cdot*_ = 1.

Supplementary Table S4 summarizes the conditions under which the distinct evolutionary outcomes identified in Fig. 1 result when both host types are subject to the same extent of isolation and quarantining. A key result is that as a generality, even modest levels of isolation can prevent the evolution and spread of the novel virus. Improved detection of symptomatic infections by the evolved virus readily causes suppression (Supplementary Table S4). Even when the ability to detect such symptomatic infections is more limited, modest isolation efforts can help limit the spread of the more asymptomatic, novel strain, with high isolation facilitating disease suppression. Still, if the ability to detect asymptomatic infections is low, there is potentially more potential for quarantining and isolation to drive infections by the novel virus. This is especially the case when there is a high transmission risk to the resilient host class, as well as a lower probability that infections by the evolved strain are asymptomatic (Supplementary Table S4 and Figs. 2A-C).

### Scenario 2: Intervention efforts reflect prevalence of symptomatic infections

As was the case for when isolation and quarantine efforts are constant through time, model (2) predicts several qualitatively distinct evolutionary outcomes when these efforts respond to the prevalence of symptomatic hosts (Fig. 3). First, above a certain isolation efficacy, most infections are with the novel strain, with the prevalence of such infections potentially depending on the quarantine efficacy (Fig 3A-3D). In contrast to a common case with constant control, when control effort tracks symptomatic infections, we find increasing public health responses can prove counter productive; higher removal efforts ultimately select for the asymptomatic strain, which subsequently spreads to high prevalence.

**Figure 3:**
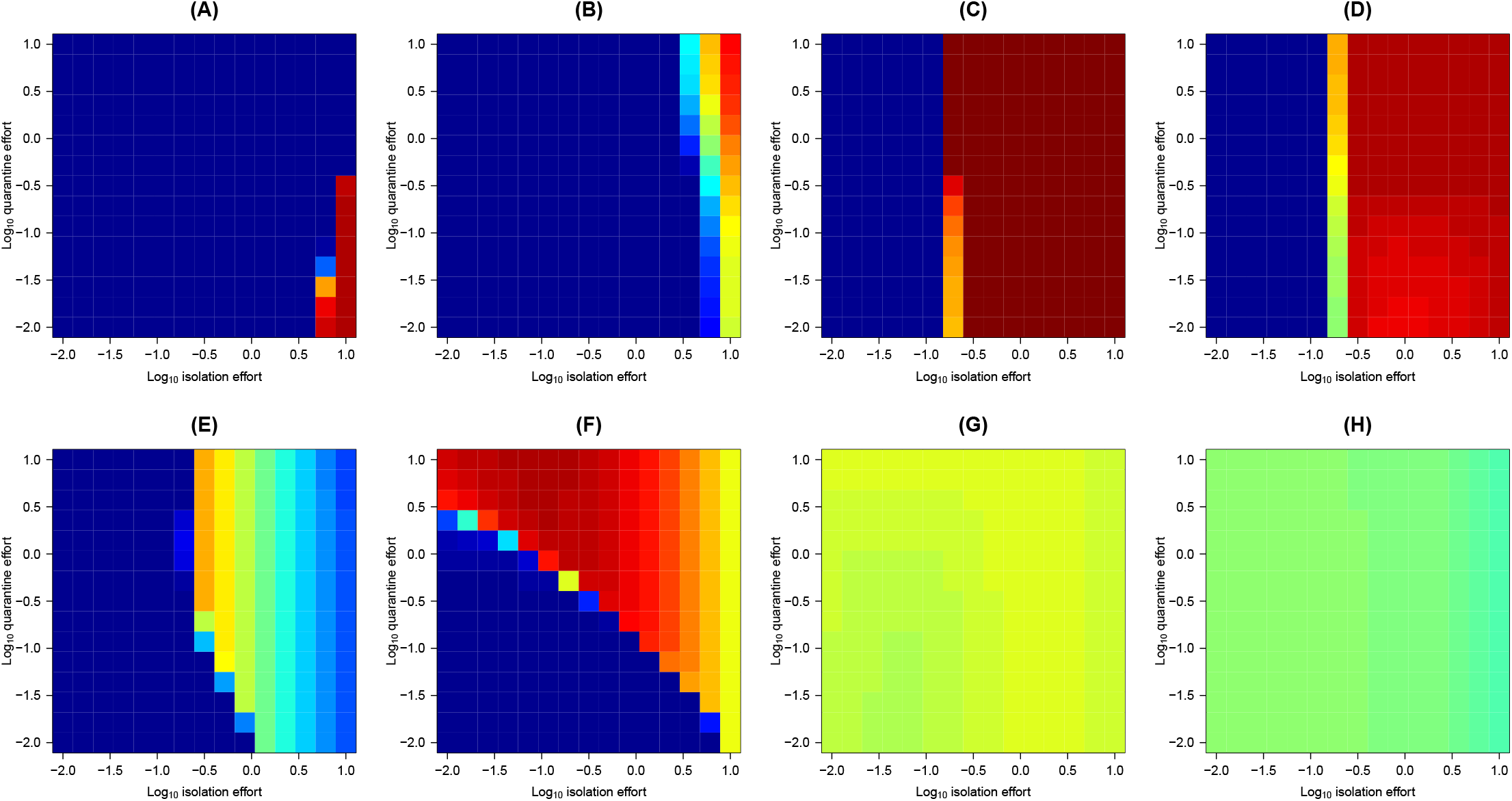
The range of possible qualitative behavior of long-term prevalence of all hosts infected with the novel, more asymptomatic virus in model (2) as a function of quarantine and isolation effort. In addition to the results above, the model also produced outcomes where the novel virus could not successfully spread or would infect all hosts irrespective of the isolation and quarantine efforts (results not shown). (A) *r*_*h,h*′_ *=* 10, *π*_1,2_*/π*_1,1_ = 1.1, *π*_2,2_*/π*_2,1_ = 1.1, *v*_*q*_ = *v*_*θ*_ = 1 and *q*_*h*,·_*/q*_*h*′,·_ = *θ*_*h*,·_*/θ*_*h*′,*cdot*_ = 0.1. (B) *r*_*h,h*′_ *=* 10, *π*_1,2_*/π*_1,1_ = 5, *π*_2,2_*/π*_2,1_ = 5, *v*_*q*_ = 0.001, *v*_*θ*_ = 1 and *q*_*h*,·_*/q*_*h*′,·_ = *θ*_*h*,·_*/θ*_*h*′,*cdot*_ = 0.1. (C) *r*_*h,h*′_ *=* 10, *π*_1,2_*/π*_1,1_ = 5, *π*_2,2_*/π*_2,1_ = 5, *v*_*q*_ = 1, *v*_*θ*_ = 0.001 and *q*_*h*,·_*/q*_*h*′,·_ = *θ*_*h*,·_*/θ*_*h*′,*cdot*_ = 0.1. (D) *r*_*h,h*′_ *=* 10, *π*_1,2_*/π*_1,1_ = 5, *π*_2,2_*/π*_2,1_ = 13, *v*_*q*_ = *v*_*θ*_ = 0.001 and *q*_*h*,·_*/q*_*h*′,·_ = *θ*_*h*,·_*/θ*_*h*′,*cdot*_ = 0.1. (E) *r*_*h,h*′_ *=* 0.1, *π*_1,2_*/π*_1,1_ = 1.1, *π*_2,2_*/π*_2,1_ = 13, *v*_*q*_ = 0.001, *v*_*θ*_ = 1 and *q*_*h*,·_*/q*_*h*′,·_ = *θ*_*h*,·_*/θ*_*h*′,*cdot*_ = 1. (F) *r*_*h,h*′_ *=* 10, *π*_1,2_*/π*_1,1_ = 1.1, *π*_2,2_*/π*_2,1_ = 1.1, *v*_*q*_ = 0.001, *v*_*θ*_ = 1 and *q*_*h*,·_*/q*_*h*′,·_ = *θ*_*h*,·_*/θ*_*h*′,*cdot*_ = 0.1. (G) *r*_*h,h*′_ *=* 1, *π*_1,2_*/π*_1,1_ = 1.1, *π*_2,2_*/π*_2,1_ = 1.1, *v*_*q*_ = 1, *v*_*θ*_ = 0.001 and *q*_*h*,·_*/q*_*h*′,·_ = *θ*_*h*,·_*/θ*_*h*′,*cdot*_ = 0.1. (H) *r*_*h,h*′_ *=* 0.1, *π*_1,2_*/π*_1,1_ = 1.1, *π*_2,2_*/π*_2,1_ = 5, *v*_*q*_ = 1, *v*_*θ*_ = 0.001 and *q*_*h*,·_*/q*_*h*′,·_ = *θ*_*h*,·_*/θ*_*h*′,*cdot*_ = 1.

A second class of possible dynamic behavior results when higher isolation efforts either facilitate suppressing the evolution and spread of the asymptomatic strain, particularly as quarantine effort decreases (Fig 3E-3F). Finally, although very subtle, in some cases modest to intermediate levels of isolation coupled with low quarantining modestly reduced the prevalence of the novel strain (Fig. 3G), while in others increasing isolation efforts are what reduced the strain’s prevalence (Fig. 3H).

Supplementary Tables S5-S6 characterize the key processes driving these qualitatively distinct eco-evolutionary predictions. A key result is that in comparison to the case where intervention efforts are constant through time, the qualitative differences in dynamical behaviors are driven much more by the detection ability of the novel strain rather than by different likelihoods of the novel virus causing asymptomatic infections.

One important result is that unlike in the case where intervention efforts are constant, high isolation efforts coupled with low quarantine efficacy most readily promote the emergence of the novel strain when the detection of asymptomatic and symptomatic infections are both high. When intervention intensity tracks the prevalence of symptomatic infections, the more symptomatic strain is quite readily removed as isolation efforts increase, and as quarantine efforts are relaxed following a reduction in the prevalence of symptomatic infections, the novel virus can then escape suppression. This outcome stands in contrast to what we found when intervention efforts were constant, in which pathogen suppression often occurred when the ability to detect asymptomatic and symptomatic infections by the evolved virus were high (Supplementary Tables S3-S4).

A somewhat different pattern emerges when most asymptomatic infections by the novel viral strain go undetected. Under this scenario, even if the detection of symptomatic infections is high, greater quarantine effort can still promote the evolution and spread of the more asymptomatic strain (Supplementary Table S5). Thus, the qualitative trend is somewhat akin to the case when intervention efforts were constant. Because quarantining efficacy is a composite of quarantining effort and the ability to detect asymptomatic cases, these results highlight how it is at intermediate levels of removing asymptomatic hosts via quarantining that results in the novel virus spreading to high prevalence. When the ability to detect and remove asymptomatic infections is high, then coupled with high isolation efforts this can facilitate disease eradication. When the ability to similarly quarantine asymptomatic infections is very low, this reduces the relative selective advantage for more asymptomatic viruses.

We further find conditions under which greater equality between host classes in public health responses diffuse selection pressure for the asymptomatic strain (Figure 4A-B, 4E and Supplementary Table S6). This contrasts to the case when the intensity of public health interventions is constant through time. The discrepancy arises because when intervention efforts track symptomatic cases, selection for being asymptomatic is low when infection rates are low. When infection rates are low, even symptomatic infections are not removed efficiently from the system. This relaxes selection pressure against the symptomatic strain. This relaxed selection pressure, in turn, prevents the asymptomatic strain from increasing to high frequency.

**Figure 4:**
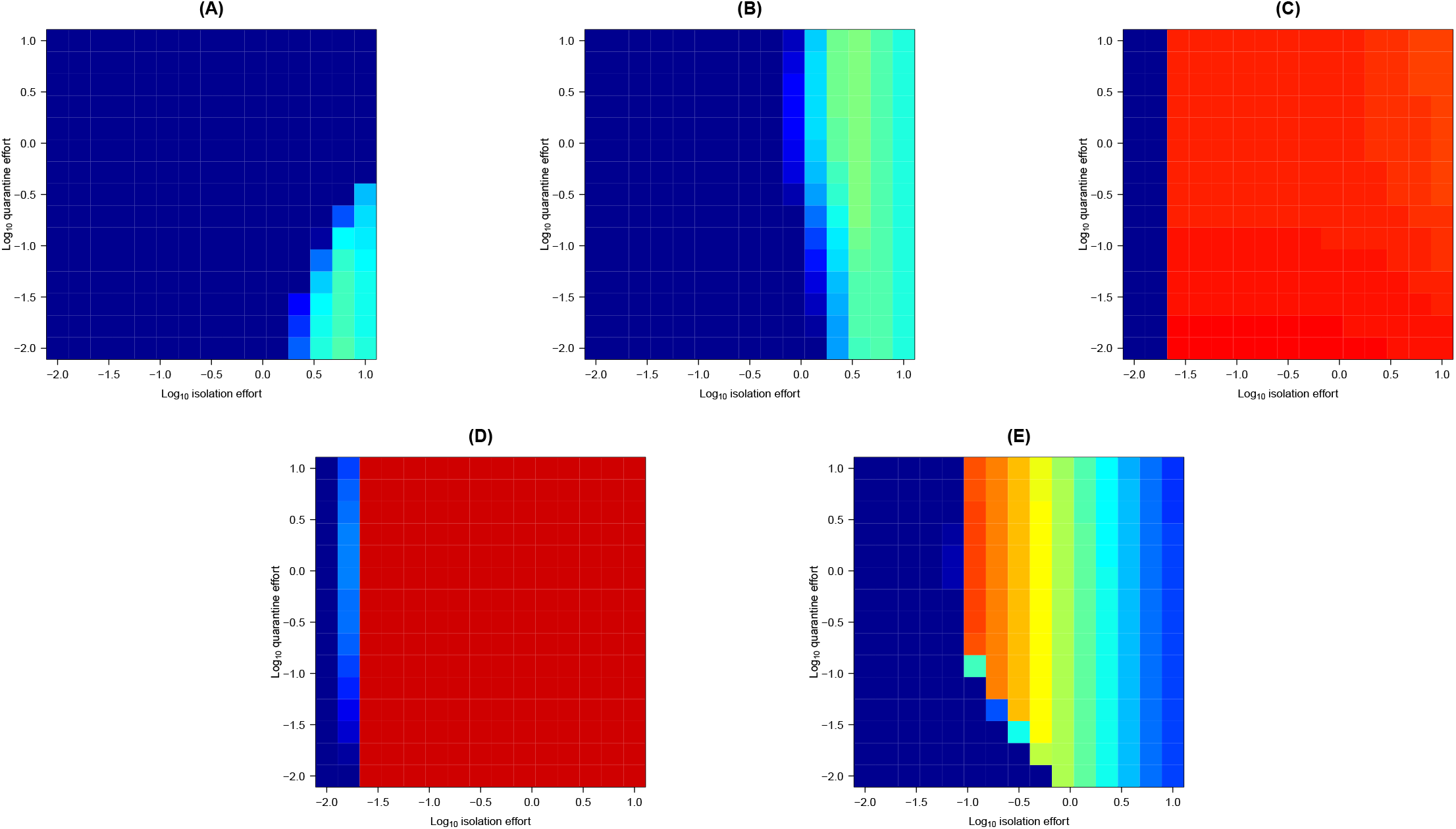
The effect of increasing isolation and quarantine efforts for the neglected host or equalizing transmission risk, in terms of long-term prevalence of among hosts infected with the novel, more asymptomatic virus in model (2) as a function of quarantine and isolation effort. (A) *r*_*h,h*′_ *=* 1, *π*_1,2_*/π*_1,1_ = 5, *π*_2,2_*/π*_2,1_ = 5, *v*_*q*_ = *v*_*θ*_ = 1 and *q*_*h*,·_*/q*_*h*′,·_ = *θ*_*h*,·_*/θ*_*h*′,*cdot*_ = 1. (B) *r*_*h,h*′_ *=* 1, *π*_1,2_*/π*_1,1_ = 5, *π*_2,2_*/π*_2,1_ = 5, *v*_*q*_ = 0.001, *v*_*θ*_ = 1 and *q*_*h*,·_*/q*_*h*′,·_ = *θ*_*h*,·_*/θ*_*h*′,*cdot*_ = 1. (C) *r*_*h,h*′_ *=* 1, *π*_1,2_*/π*_1,1_ = 5, *π*_2,2_*/π*_2,1_ = 5, *v*_*q*_ = 0.001, *v*_*θ*_ = 1 and *q*_*h*,·_*/q*_*h*′,·_ = *θ*_*h*,·_*/θ*_*h*′,*cdot*_ = 1. (D) *r*_*h,h*′_ *=* 10, *π*_1,2_*/π*_1,1_ = 5, *π*_2,2_*/π*_2,1_ = 5, *v*_*q*_ = 0.001, *v*_*θ*_ = 0.001 and *q*_*h*,·_*/q*_*h*′,·_ = *θ*_*h*,·_*/θ*_*h*′,*cdot*_ = 1. (E) *r*_*h,h*′_ *=* 100, *π*_1,2_*/π*_1,1_ = 1.1, *π*_2,2_*/π*_2,1_ = 5, *v*_*q*_ = 0.001, *v*_*θ*_ = 0.001 and *q*_*h*,·_*/q*_*h*′,·_ = *θ*_*h*,·_*/θ*_*h*′,*cdot*_ = 0.11.

At the same time, in some cases even quite low isolation efforts can induce the evolution and spread of the more asymptomatic virus, with quarantine effort having little effect (Fig. 4C-D). This outcome mirrors the results for the constant control effort case characterized in, e.g., Fig. (2I). We find this to especially be the case if most infections are of the more common, vulnerable host class that experiences lower isolation and quarantine efficacy. When the intensity of public health intervention is even, a higher infection risk for the vulnerable host class can amplify selection pressure for the more asymptomatic strain. Indeed, increasing the ability to detect infections of the vulnerable host class selects for the novel virus when there is enhanced infection risk among the vulnerable host class (Supplementary Table S6). When infection risks are more even across host classes, improving isolation and quarantine efforts for the vulnerable host class reduces selection for the more asymptomatic strain (Supplementary Table S6).

## Discussion

Because many emerging zoonotic pathogens lack effective prophylactics or treatment, the identification and removal of infectious individuals remains the primary control strategy (Eames and Keeling 2003; Bird and Mazet 2018). When epidemiological monitoring fails to detect asymptomatic carriers, the pathogens they harbor are able to spread with less friction through the host population. By contrast, potentially symptomatic lineages are subject to detection and, through contact-tracing even asymptomatic infections are identified quarantined, thereby pruning that particular viral lineage.

Our model analyses using SARS-Coronoviruses as a case study illustrates how reliance on symptoms-driven reporting and control, however well-meaning, can ultimately shift the pathogen’s fitness landscape to select for pandemic strains. We show that such selection for an asymptomatic pathogen is often most acute when isolation and quarantine efforts are intense, but fall short of complete disease suppression. Our comparative analyses further indicate that when host removal depends on the prevalence of symptomatic infections, even very high levels of isolation effort can facilitate the emergence and extensive spread of more asymptomatic strains.

One implication is that although most public health responses to emerging zoonotic pathogens are necessarily reactive, an anticipatory approach to case identification may be necessary for highly infectious pathogens such as coronaviruses known to cause asymptomatic infections. For instance, routine, widespread randomized metagenomic surveys of known and suspected reservoirs would more closely approximate the “constant intervention efforts” scenario we model. Our results indicate, however, that such efforts need to be quite effective to prevent the emergence of asymptomatic viruses. Whether anticipatory control measures such as these are realistic given constraints on public health budgets remains an open question (Waitzkin 2018).

We further highlighted the critical role host heterogeneity plays in driving the evolutionary consequences of isolation and quarantining symptomatic individuals. By definition, emerging zoonotic pathogens navigate a heterogenous landscape of hosts, and even after evolving to circulate in humans, social differences result in distinct intervention efficacies and transmission chains (Levins 1995; Szreter and Woolcock 2004; Wallace et al. 2015; Dzingirai et al. 2017a). These differences imply that selection pressures experienced by pathogens is variable, and our results show how such variable selection can promote the evolution and spread of more asymptomatic viruses.

We find in general that implementing isolation and quarantine evenly across host classes can facilitate disease suppression and thereby reduce selection for more asymptomatic viruses. However, reducing disparities among host classes can also increase overall selection pressure against more symptomatic strains in some cases. For instance, when isolation and quarantine efficacy just low enough to permit the suppression of a more asymptomatic virus by a less asymptomatic virus, evenly carrying out isolation or quarantining efforts across host classes can occasionally shift the balance to promote the spread of the more asymptomatic virus.

One dimension of host heterogeneity that had a slightly counter-intuitive effect was when transmission risk was more even across host classes, this in fact promoted suppression. We found that at least in some cases, increasing transmission for the resilient host operates, to some degree, in a manner analogous to the dilution effect, in which viral infections of hosts that cannot transmit the pathogen slow the epidemic (e.g., Roberts and Heesterbeek 2018). When resilient hosts are likelier to be successfully identified and removed from the population of infectious hosts, the ability of isolation and quarantine to suppress the disease becomes more apparent when these hosts are more likely to get infected. One consequence of this is that when resilient hosts, who are likelier to be successfully identified and removed if infected, are less likely to be exposed to the pathogen, then this can create conditions conducive the evolution and spread of a more asymptomatic strain. Thus, evening transmission risk across host classes may be one strategy for disease control when infections in some hosts are likelier to be removed from the population.

By design, but also due to limitations in the data, our numerical analyses surveyed a wide segment of parameter space. Given the frequency with which a more asymptomatic coronavirus emerged and spread in our simulations, as well as the distinct qualitative outcomes of the role of isolation and quarantine efforts, we highlight the need for better measurements of key quantities. A case in point is the degree to which mutations enable viruses to cause asymptomatic infections. Because selection against symptomatic strains involves a reciprocal interplay between epidemiological and evolutionary processes, the evolution and spread of the novel strain at times depended on the probability that infection with the novel virus is asymptomatic. Thus, any intervention measure aimed at preventing the emergence of an asymptomatic strain must take into account the viral genotype-phenotype map. To be sure, whether a virus causes symptoms also depends heavily on the host’s biology. Nevertheless, in light of our results, given the growing capacity to characterize large amounts of viral sequence variation, explaining how this variability drives the propensity of viral infections to be symptomatic (such as Korber et al. 2020) seems particularly warranted.

In conclusion, our analyses show how the evolution and spread of asymptomatic viruses is driven by a reciprocal interplay between public health intervention measures and prevailing host population structure, on the one hand, and the nature of the host-pathogen interaction at the level of individual hosts on the other. We think our results also have implications for endemic pathogens. Several other major human pathogens (dengue, Methicillin-resistant Staphylococcus aureus) similarly lack effective and widespread prophylactics or treatment. In light of our conclusions, we urge epidemiological monitoring efforts to seriously consider undertaking randomized testing to avoid inadvertently creating a surveillance regime that selects for more asymptomatic strains, and to do so in a way that is consistent across host classes. Our investigation provides a critical first step towards providing quantitative guidance for determining evolutionarily appropriate levels of interventions for coronaviruses and other highly transmissible pathogens.

## Data Availability

This is theoretical work so there is no data. Interested readers are invited to consult https://github.com/kewok/AsymptomaticEvolution for code used in the analyses

## Acknowledgments

The authors would like to thank P. Turner for valuable comments on an earlier version of this manuscript.

## Supplementary Material

Supplementary Material S1 Showing little effect of varying *δ* from 0.05 to 0.85 with constant intervention effort

Supplementary Material S2 Showing little effect of varying *δ* from 0.05 to 0.85 with variable intervention effort

Supplementary Tables S3-S4. A description of the effects of varying the parameter values pertaining to host heterogeneity and viral phenotype on the qualitative outcomes under varying intervention efforts. The qualitative behavior for parameter combinations are categorized according to the panels depicted in Fig. 1, when the isolation and quarantine efforts differ across host types (Supplementary Table S3) and when they are concomitant across host types (Supplementary Table S4). (*) denotes instances where the novel strain did not spread or where the pathogen was eradicated regardless of infectious host removal efforts, (**) denotes instances where the novel strain did not spread, but the pathogen persisted (often at very low prevalence) regardless of infectious host removal efforts, and (X) denotes instances where the novel pathogen spread throught the entire host population regardless of infectious host removal efforts.

Supplementary Tables S5-S6. A description of the effects of varying the parameter values pertaining to host heterogeneity and viral phenotype on the qualitative outcomes under varying intervention efforts. The qualitative behavior for parameter combinations are categorized according to the panels depicted in Fig. 3, when the isolation and quarantine efforts differ across host types (Supplementary Table S5) and when they are concomitant across host types (Supplementary Table S6). As in Supplementary Tables S3-S4, (*) denotes instances where the novel strain did not spread or where the pathogen was eradicated regardless of infectious host removal efforts, (**) denotes instances where the novel strain did not spread, but the pathogen persisted (often at very low prevalence) regardless of infectious host removal efforts, and (X) denotes instances where the novel pathogen spread throught the entire host population regardless of infectious host removal efforts.

