## Supplementary material for "When might Host Heterogeneity Drive the Evolution of Asymptomatic, Pandemic Coronaviruses?": SupplementaryResults1.txt

Percentiles for difference in prevalence of strain 2 between when delta=0.05 and delta=0.85 when the intervention effort is constant across all parameter combinations

            1%             5%            15%            25%            35% 
 -5.322333e-06  -1.554312e-15  -1.110223e-16  -2.032879e-20 -6.258217e-264 
           45%            55%            65%            75%            85% 
 3.063608e-108   6.899117e-27   7.453890e-20   2.220446e-16   1.811332e-13 
           95%            99% 
  4.697390e-05   3.734351e-03 
