## Supplementary material for "When might Host Heterogeneity Drive the Evolution of Asymptomatic, Pandemic Coronaviruses?": SupplementaryResults2.txt

Percentiles for difference in prevalence of strain 2 between when delta=0.05 and delta=0.85 when the intervention effort responds to infection prevelance across all parameter combinations 

          1%            5%           15%           25%           35% 
-7.671014e-02 -1.781497e-02 -4.181629e-04 -4.102562e-06 -5.194552e-08 
          45%           55%           65%           75%           85% 
-4.996004e-16 -1.084202e-19 -1.694066e-21  1.355253e-20  3.903128e-18 
          95%           99% 
 4.013999e-08  3.254309e-03 
